# Intradiscal propionic acid levels in chronic low back pain patients with Modic type 1 changes are associated with systemic immune cell activation

**DOI:** 10.64898/2026.01.06.26343445

**Authors:** Jan Devan, Jiamin Zhou, Po-Hung Wu, Noah Bonnheim, Conor O’Neill, Jeffrey C. Lotz, Thomas M. Link, Abel Torres-Espin, Stefan Dudli, Aaron J. Fields, the REACH Center at UCSF

## Abstract

**Background:** Modic type 1 changes (MC1) are vertebral endplate bone marrow lesions strongly associated with chronic low back pain (CLBP), but their underlying pathobiology remains unclear. Recent findings suggest that disc–bone marrow crosstalk that includes *Cutibacterium acnes (C. acnes)* infection of the disc, and immune system activation in the adjacent vertebrae may play a significant role.

**Methods:** In a prospective analysis of patients from the UCSF comeBACK cohort, we quantified intradiscal propionic acid (PA) – a metabolic product of *C. acnes* – using magnetic resonance spectroscopy (MRS). Patients were stratified into tertiles based on intradiscal PA content, and the uppermost (PA-high) and lowest (PA-low) tertiles were compared for systemic immune signatures, including whole-blood transcriptomics (n=196), flow cytometry immunophenotyping (n=224), and serum cytokine profiling (n=398).

**Results:** Elevated intradiscal PA was associated with distinct systemic immune responses in patients with MC1 but not in patients with Modic type 2 or without Modic changes. Transcriptomic analysis revealed enrichment of adaptive immune pathways and B cell activation signatures in PA-high MC1. Flow cytometry identified the expansion of immunosuppressive ectonucleotidases-expressing B cells in PA-high MC1 patients. Finally, the correlation profiles of intradiscal PA levels and circulating B cells with serum cytokine concentrations were highly similar. Together, these data consistently indicate that systemic B cell activation is a hallmark of PA-high MC1.

**Conclusion:** Intradiscal PA, measurable non-invasively by MRS, stratifies MC1 patients into biologically distinct subgroups characterised by systemic B cell activation. These findings suggest a systemic immune component to MC1 pathobiology and highlight B cells as candidate biomarkers and therapeutic targets for MC1-related chronic low back pain.

## 1. INTRODUCTION

Low back pain (LBP) is the most common musculoskeletal condition and is the leading cause of disability worldwide. The aetiology of LBP is multifactorial^1^. In some patients, LBP may be caused by spinal pathologies that appear on magnetic resonance imaging (MRI). In particular, vertebral endplate bone marrow lesions, which appear as signal intensity changes on clinical T1-weighted and T2-weighted MRI (“Modic changes”, or MC), are significantly associated with chronic LBP (CLBP)^2,3^ and have a higher prevalence in LBP patients than in asymptomatic controls^4^. Clinical management of CLBP in patients with MC is challenging because treatment outcomes are heterogeneous in this subgroup ^5^. The heterogeneity in outcomes may reflect differences in MC pathophysiology and/or aetiology that are presently indistinguishable using conventional MRI sequences alone^6,7^. Differences in MC pathophysiology could involve differences in bone marrow activation, differences in blood immune cell composition, and differences in cytokine profile^8–11^. Identifying differences in the blood immune profile of CLBP patients with MC could, therefore, clarify MC pathophysiology, motivate diagnostic approaches that allow for better patient stratification, and inform therapeutic strategies that more precisely target the underlying pathophysiology.

Prior research indicates that MCs coincide with elevated disc-vertebra crosstalk at sites of endplate damage^12^ and that the pathomechanisms of MC bone marrow contain autoimmune and innate immune responses^10,13,14^. Endplate damage predisposes to MC and exposes bone marrow immune cells to disc tissues and tissue-secreted factors ^6^. Involvement of the adaptive immune system in MC pathobiology is evidenced by our discovery that MC significantly associates with HLA-alleles^15^. Separately, some MC-adjacent discs have high numbers of *Cutibacterium acnes (C. acnes)*, a bacterium that can cause MC in animal models^16^. In human MC-adjacent discs, high numbers of *C. acnes* have been associated with the activation and accumulation of neutrophils^11^.

To understand the pathobiology of MC, prior studies focused on biopsies collected intraoperatively during spinal fusion surgery^9,12,13^, which is the gold standard for profiling the local cell and molecular milieu. However, because this sampling procedure is limited to surgical patients, the practical implications for patient stratification in future studies are unclear. Likewise, because the sampling procedure is a potential source of biopsy contamination, the clinical validity of the findings is a matter of controversy. We address these issues in a large cohort of CLBP patients using a non-invasive methodology sensitive to intradiscal *C. acnes*^17^. Specifically, we utilised measures of intradiscal propionic acid (PA), a metabolite of *C. acnes*, obtained from disc magnetic resonance spectroscopy (MRS) to stratify patients into high and low PA groups, and we compared the blood transcriptome, immune cell populations, and cytokine profiles between these patient groups. Using this approach, we sought to test the hypothesis that patients with high and low intradiscal PA levels differ in their blood immune profile, enabling non-invasive stratification of MC subtypes.

## 2. METHODS

This prospective study was conducted with institutional review board approval (WIRB #20-30368) and consent of the cantonal ethics commission (BASEC #2019-00845). Written informed consent was taken from all individual participants.

### 2.1. Subjects

A subset of participants in The Longitudinal Clinical Cohort for Comprehensive Deep Phenotyping of Chronic Low-Back Pain Adults Study (comeBACK)^18^, a part of the NIH Back Pain Consortium (BACPAC)^19^, were included in this analysis. To be eligible for inclusion in the present study, comeBACK participants must have had (a) an available clinical MRI and single-voxel MRS acquisitions and (b) blood serum and blood RNA samples collected at the time of MRI and MRS. Of the 463 total participants enrolled in comeBACK, 398 patients had blood sampled at their baseline visit available for cytokine quantification and 389 for total blood transcriptomic analysis. Of those 398 patients, 200 patients underwent baseline MRI and MRS. All participants had LBP for at least three months and at least half of the days in the past 6 months.

### 2.2. Imaging

Lumbar spine MRI was performed on a Discovery MR 750 3-T scanner using an 8-channel phased-array spine coil (GE Healthcare). Clinical fast spin-echo images with T_1_ and T_2_ weighting were acquired using sagittal and axial prescriptions, as detailed previously^20^. MC grading was performed by a musculoskeletal radiologist on T_1_- and fat-saturated T_2_-weighted images according to standard MC definitions: type 1 MC (MC1) were hypointense on T_1_- and hyperintense on fat-sat T_2_ images; type 2 MC (MC2) were hyperintense on T_1_- and hypointense on fat-sat T_2_-weighted images. Patients presenting with both MC1 and MC2 at different lumbar levels or within the same endplate (so-called “mixed” MC) were categorised as MC1, reflecting the more clinically significant MC with increased inflammation and association with pain^21,22^.

### 2.3. Magnetic resonance spectroscopy

Single-voxel MRS spectra were acquired on a Siemens Skyra 3-T scanner using a Point-RESolved Spectroscopy (PRESS) sequence and Chemical Selective Suppression (CHESS) for water suppression ^23^. Briefly, shortened T_1_-weighted and T_2_-weighted acquisitions in the sagittal, coronal, and axial planes enabled voxel prescription to encompass the disc nucleus pulposus while excluding the vertebral body. Shimming was performed to optimise the water signal before initiating the MRS acquisition series of 160-192 frames using a 16-step phase cycle (1500 ms TR, 32 ms TE). Disc levels for MRS were selected by the scanner operator, who chose up to four lumbar levels per patient and acquired the spectra in a caudal-to-cranial order, blinded to the radiologist’s MC diagnoses. Acquired spectra were post-processed using the NOCISCAN-LS™ software (Aclarion, Inc.; Broomfield, CO), which optimised spectral quality, including sufficient signal-to-noise ratio, frame editing, phase and frequency shift error correction, baseline correction, and artefact correction. The area under the curve (AUC) of the PA peak (chemical shift ∼1.05 ppm) was normalised to voxel size. The maximum PA of all discs analysed per patient was then used to stratify patients into PA tertiles: “PA-low” (little or no intradiscal bacteria present); “intermediate PA” (uncertain bacterial presence); and “PA-high” (higher amount of intradiscal bacteria present).

### 2.4. Blood sample collection

Whole blood samples from patients with varying fasting status were collected in the morning after imaging acquisition in one 10 mL serum separating tube for serum analysis and one 2.5 mL PAXgene blood RNA tube for RNA analysis^24^. Serum samples were allowed to clot at room temperature for 30 minutes and then centrifuged for 15 minutes at 1300 rcf at 4°C. The resulting serum supernatant was aliquoted into labelled cryovials and frozen at −80°C for storage. For RNA PAXgene analysis, the PAXgene blood RNA tubes were stored upright at room temperature for 2 hours, frozen at −20°C for 24 hours, and then transferred and stored at −80°C.

### 2.5. Transcriptomics

RNA was isolated from PAXgene Blood RNA tubes using RNeasy Mini Kit (Qiagen) and QiaCube Connect (Qiagen). RNA libraries were prepared for each sample with RNA Integrity Number ≥ 6 and RNA concentration > 5 ng/μL using TruSeq Stranded mRNA (Illumina). The cDNA libraries were sequenced using 100 cycles on a NovaSeq X platform (Illumina) with 100bp single reads to an average of 26M reads. Data were processed using SUSHI, a fully documented and reproducible pipeline for RNA sequencing data from raw data to differential expression analysis^25,26^. This includes *FastQC* to control the quality of reads, Kallisto for genome alignment and quantification of transcript abundance, DESeq2 and clusterProfiler for Gene Set Enrichment Analysis (GSEA)^27,28^.

For blood transcriptomics, Principal Component Analysis (PCA) of the top 2000 features were performed to show unbiased population-specific clustering and provide a systematic view of gene differential expression. Significantly differentially expressed genes (DEG) were determined as p<0.01 and log2 fold-change>0.5. A False Discovery Rate (FDR) ≤0.05 was used to detect candidate terms in GSEA.

### 2.6. Flow cytometry immunophenotyping of blood cells

Cells isolated by erythrocyte lysis and centrifugation of peripheral blood were cryopreserved. After thawing, they were incubated for 1 day in Roswell Park Memorial Institute (RPMI) medium supplemented with 10% heat-inactivated fetal calf serum, penicillin/streptomycin, and stable glutamine. Afterwards, cells were stained with an immunophenotyping panel (**Tab. S1**), fixed with fixation buffer (Biolegend) and acquired on a cytometer (Cytek Aurora, five lasers). All samples were stained and analysed in one experiment. Unmixing of spectral flow cytometry data was done using SpectroFlo (v2.3; Cytek Biosciences), compensations and gating were performed in FlowJo (v10.10; BD Biosciences), PCA analysis, 2-way ANOVA and correlation analyzes were done GraphPad Prism, FlowSOM clustering was performed in R (v4.3.2)^29^. Comparison of PC scores or proportions of specific immune cells between patient groups defined by PA levels was done using ordinary one-way ANOVA or Brown-Forsythe and Welsch ANOVA test based on the results of Bartlett’s test for differences in standard deviations (SD).

### 2.7. Serum cytokine measurement

Serum samples were assayed for simultaneous quantitative measurement of 40 inflammation-associated cytokines using Quantibody® Human Inflammation Array 3 (QAH-INF-3; Raybiotech, Inc.; Norcross, GA), an array-based multiplex sandwich enzyme-linked immunosorbent assay (ELISA). Missing cytokine concentrations were imputed using principal component analysis (PCA) imputation, which estimates missing values based on PCA while minimising the influence of outliers. Results were filtered to exclude values below the limit of detection (LOD) and above the maximum for each cytokine (**Tab. S2**). Afterwards, serum cytokine concentrations were log-transformed to reduce the effects of skewness in the distributions.

Statistical analysis was performed using GraphPad Prism. Spearman correlation coefficients between cytokine concentrations and PA, and between cytokine concentrations and proportions of immune cell subsets, were calculated, and hierarchical agglomerative clustering using Euclidean distance and complete linkage was used to investigate similarities in the correlation coefficients of immune cell subsets and intradiscal PA levels with blood plasma cytokine concentrations.

## 3. RESULTS

### Patient characteristics

The analysis included 398 CLBP patients from the comeBACK study with available MRS. For 200 of these patients, lumbar spine MRI data were available, allowing diagnosis of the Modic changes. The average age of these 200 patients was 51.3 ± 15.6 years, and 42.5% were male (**Tab. 1**). Patients reported an average pain rating in the week before study participation of 4.5 ± 1.9 on the Pain, Enjoyment of Life and General Activity (PEG) scale, which ranges from no pain (0) to pain as bad as you can imagine (10). On the Patient-Reported Outcomes Measurement Information System (PROMIS) assessment of pain interference (PI) and physical function (PF), patients reported an average T-score of 58.5 ± 7.7 in PI and 41.8 ± 6.4 in PF, corresponding to higher average pain interference and lower average physical function than the reference general population from the United States.

**Table 1.** Demographics and clinical characteristics of LBP patients.

| Characteristic | Total<br>(*n* = 200) | Modic change |  |  | *p*-value |
| --- | --- | --- | --- | --- | --- |
|  |  | MC0<br>(*n* = 85) | MC1<br>(*n* = 80) | MC2<br>(*n* = 35) |  |
| Demographics |  |  |  |  |  |
| Age (year) | 51.3 ± 15.6 | 45.8 ± 16.8<sup>a</sup> | 54.5 ± 13.4<sup>b</sup> | 57.2 ± 13.0<sup>b</sup> | <0.0001<sup>***</sup> |
| Male | 85 (42.5) | 31 (36.5) | 39 (48.8) | 15 (42.9) | 0.39 |
| BMI (kg/m<sup>2</sup>) | 26.2 ± 4.7 | 25.4 ± 4.1<sup>a</sup> | 26.0 ± 3.9<sup>a</sup> | 28.5 ± 6.5<sup>b</sup> | 0.003<sup>**</sup> |
| Clinical measures |  |  |  |  |  |
| PEG | 4.5 ± 1.9 | 4.4 ± 1.8 | 4.6 ± 1.9 | 4.4 ± 1.7 | 0.84 |
| PROMIS PI | 58.5 ± 7.7 | 58.6 ± 6.8 | 57.8 ± 8.3 | 59.9 ± 8.4 | 0.42 |
| PROMIS PF | 41.8 ± 6.4 | 41.7 ± 5.6 | 42.3 ± 6.9 | 40.9 ± 7.0 | 0.56 |
| PA | 3.2 ± 3.2 | 3.2 ± 3.0 | 3.1 ± 3.4 | 3.4 ± 3.1 | 0.86 |

<sup>\*\*</sup> $p < 0.01$ , <sup>\*\*\*</sup> $p < 0.001$
For continuous numerical variables, mean values ± standard deviations are indicated. Overall group differences between MC types were tested with one-way ANOVA. For count data (Male), the count is indicated, and the percentage is in parentheses. Pairwise comparisons using Tukey-Kramer HSD were performed for asterisked characteristics, and connecting groups are marked with superscripts (a, b); all have a significance of $p < 0.05$ . BMI: body mass index; PEG: pain scale (0-10); PROMIS PI: Patient-Reported Outcomes Measurement Information System pain interference T-score; PROMIS PF: physical function T-score; PA: maximum measured intradiscal PA.

Eighty-five of the 200 patients (42.5%) had no MC (MC0) present at any lumbar level, 80 (40%) had at least one lumbar level graded as MC1, and 35 (17.5%) had only MC2 graded along their lumbar spine. There were no significant differences in sex or clinical measures between patients in each MC classification. Age was significantly lower in MC0 patients compared to either MC1 or MC2 patients (45.8 years *vs.* 54.5 and 57.2 years, respectively; *p* < 0.0001), and BMI was significantly higher in patients with only MC2 compared to MC0 or MC1 (28.5 kg/m^2^ *vs.* 25.4 and 26.0 kg/m^2^, respectively; *p* = 0.003).

### Measurement of intradiscal propionic acid

Intradiscal PA levels were measured with MRS in 530 total lumbar discs from 200 patients (1-4 discs/patient). The distribution of patients’ maximum PA levels ranged from 0.00 to 18.61. The maximum PA level in each patient’s lumbar spine was 3.2 ± 3.2, with a positive skewness of 2.0 and a positive kurtosis of 5.3. The maximum PA level was used to stratify patients into tertiles. Patients with “PA-low” (*n* = 69) had PA levels below 1.57, and patients with “PA-high” (*n* = 63) had PA levels above 3.65. All PA groups had similar age, sex, and BMI. There was no significant difference in intradiscal PA levels between MC1, MC2 and MC0 patients (**Fig. S2**).

### Transcriptional signature of adaptive immune response in “PA-high” of MC1

Total blood RNA was sequenced from 196 patients; 4 did not pass the quality threshold. Eighty-four had MC1 (PA-low: 28, PA-high:27), 34 had MC2 (PA-low: 12, PA-high: 11), and 78 had MC0 (PA-low: 25, PA-high: 27). The first 10 PC of the 2000 most variable genes explained 44.6% of total data variance (**Fig.S1A**). Samples did not cluster according to MC type (**Fig.S1B**) or PA level (**Fig.S1C**). Comparing PA-high vs PA-low in MC1, 21 genes were up- and 13 downregulated, in MC2, 46 were up-and 15 downregulated, and in MC0, 55 were up- and 20 downregulated (**Fig. 1A, B**). No gene was consistently up- or down-regulated in PA-high vs PA-low in all MC types. Immunoglobulin Heavy Constant Gamma (IGHG) 3 and Interferon Alpha Inducible Protein 27 (IFI27) were both upregulated in the PA-high group of MC1 and MC0. IGHG1 was upregulated in the PA-high group of MC1 and MC2. GSEA revealed enrichment of biological processes related to adaptive immune response in PA-high vs PA-low in MC1 and MC2, but not in MC0 (**Tab. 2**). Activation of B cells, indicated by the GO terms “Positive regulation of B cell activation” and “B cell receptor signaling pathway” were only enriched in MC1, but not in “PA-high” of MC0 and MC2. Furthermore, innate immune processes linked to antibacterial defense mechanisms were enriched in “PA-high” vs “PA-low” in MC1 and MC0, but not in MC2 (**Tab. 2**). The gene set “defense response to bacteria” contained many immunoglobulins heavy chain transcripts that were shared with the gene set “adaptive immune response” and are involved in “phagocytosis, recognition” (**Fig. 1C**). Of the remaining 32 genes, 17 were part of the gene set “Neutrophil degranulation” and were overrepresented in the GO terms “Secretory Granule Lumen” (overlap: 15/316, *p*-value=1.2E-17) and “Azurophil Granule Lumen” (overlap: 8/89, *p*-value=2.1E-11), suggesting neutrophil activation. The differential expression of these neutrophil-related genes between ‘PA-high’ and ‘PA-low’ conditions distinguished MC1 from both MC2 and MC0 (**Fig. 1D**).

**Figure 1.**
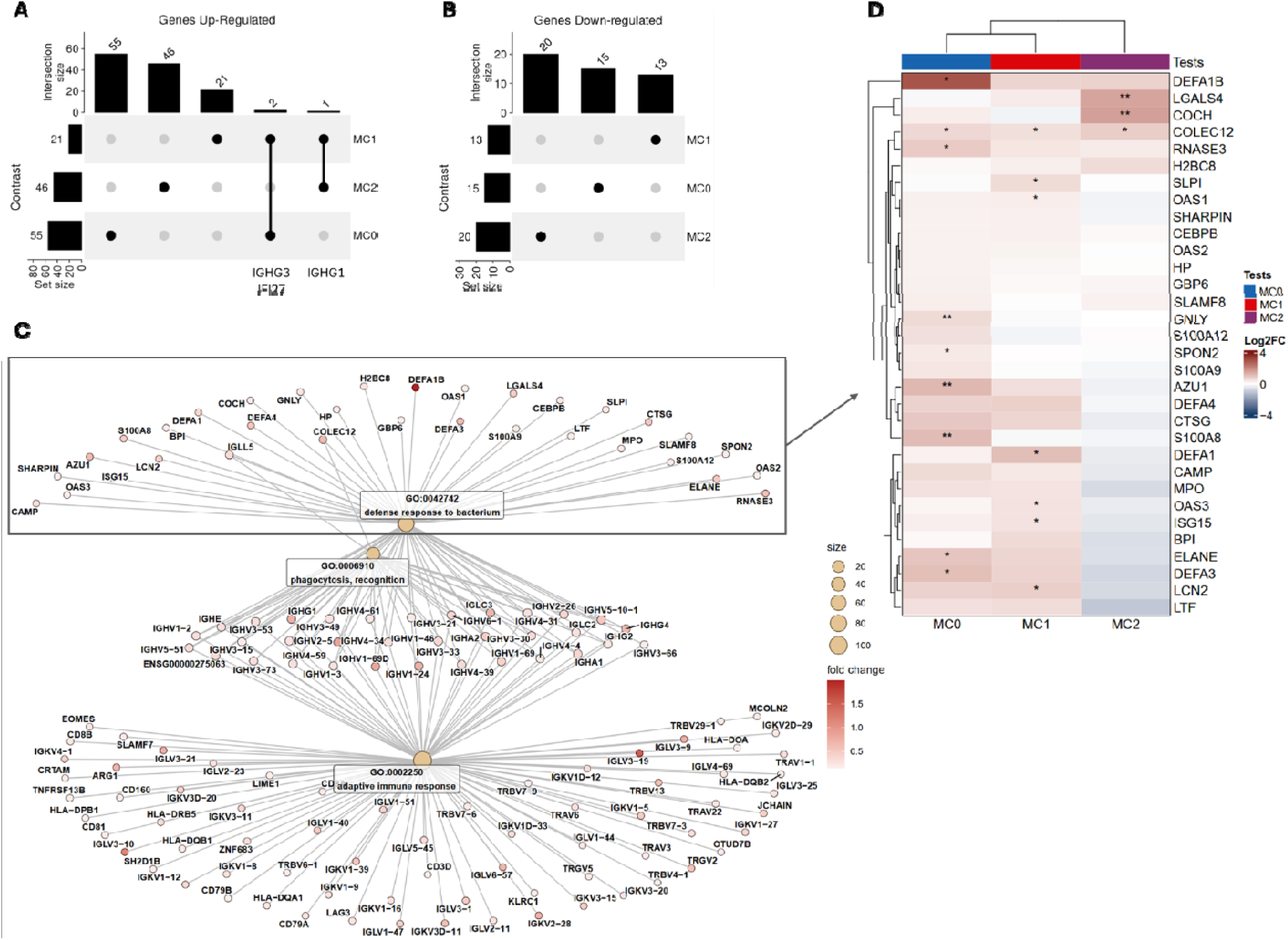
Whole blood bulk-RNA sequencing analysis from CLBP patients. (**A**) Upset plot intersecting up-regulated genes in patients with “PA-high” vs. “PA-low” between MC types. Gene names are given for intersections below the plot. (**B**) Upset plot intersecting down-regulated genes in patients with “PA-high” vs. “PA-low” between MC types. (**C**) Network plot of GO terms “Defence response to bacterium” and “Adaptive immune response”. Node size indicates the count of genes. The dot colour of gene indicates fold-change upregulation. Rectangle indicates genes analyzed in subfigure D. (D) Heatmap showing genes from the GO term “Defense response to bacterium” that were not shared with “Adaptive immune response”. Red colour indicates upregulation, blue colour downregulation. Hierarchical clustering of MC type is indicated at the top, and hierarchical clustering of genes on the left side. Asterisks indicate significance of up-/downregulation: * p<0.05, ** p<0.01.

**Table 2.** Whole blood bulk-RNA sequencing analysis from CLBP patients. Enrichment of gene sets in “PA-high” vs. “PA-low” by Modic change (MC) type.

| GO ID | Description | MC1 |  | MC2 |  | MC0 |  |
| --- | --- | --- | --- | --- | --- | --- | --- |
|  |  | NES | p-value | NES | p-value | NES | p-value |
| Adaptive immune response |  |  |  |  |  |  |  |
| GO:0002250 | Adaptive immune response | 1.85 | <0.0001*** | 1.90 | 0.0001*** |  |  |
| GO:0050871 | Positive regulation of B cell activation | 2.04 | 0.0014** |  |  |  |  |
| GO:0050853 | B cell receptor signalling pathway | 1.89 | 0.0182* |  |  |  |  |
| Innate immune response |  |  |  |  |  |  |  |
| GO:0042742 | Defence response to bacterium | 2.00 | 0.0003*** |  |  | 1.87 | 0.0022** |
| GO:0006910 | Phagocytosis, recognition | 2.09 | 0.0007*** |  |  |  |  |
| GO:0006911 | Phagocytosis, engulfment | 1.94 | 0.0185* |  |  |  |  |
| GO:0050829 | Defence response to Gram-neg.bacterium |  |  |  |  | 1.87 | 0.0384* |
| GO:0019731 | Antibacterial humoral response |  |  |  |  | 1.91 | 0.0262* |
| GO:0050832 | Defence response to fungus |  |  |  |  | 1.92 | 0.0262* |
| Protein translation |  |  |  |  |  |  |  |
| GO:0002181 | Cytoplasmic translation |  |  |  |  | 2.14 | <0.0001*** |
| GO:0006412 | Translation |  |  |  |  | 1.70 | 0.0022** |

<sup>\*</sup> $p < 0.05$ , <sup>\*\*</sup> $p < 0.01$ , <sup>\*\*\*</sup> $p < 0.001$

### Immunophenotyping by flow cytometry

Peripheral blood cells from 224 patients with CLBP were immunophenotyped using multicolour flow cytometry. Both MRS and MRI were accessible for 170 of them. The marker panel distinguished 45 main immune cell lineages and 1021 smaller cell populations corresponding to different activation or functional maturation stages of the main lineages. Samples from patients with available MC classification and intradiscal PA levels were further analysed: 73 MC1 (PA-low: 26; intermediate PA: 26; PA-high: 21), 29 MC2 (PA-low: 11; intermediate PA: 11; PA-high: 9), and 68 MC0 (PA-low: 23; intermediate PA: 22; PA-high: 23).

PCA was used to reduce the dimensionality of the flow cytometry data (**Fig. 2A**). The first eight PCs cumulatively explained 68.7% of the variance in cell subset percentages, with PC1 capturing 21.8%, PC2 10.0%, and PC3 8.8% (**Fig. 2A, B**). When comparing PC values between patients with PA-low (PA-low) and PA-high (PA-high), PC3 values were significantly higher in patients with MC1 and PA-high (PA-high) than in those with MC1 and PA-low (PA-low) (**Fig. 2B**). PC3 values were weighted heavily by B cell signature (**Fig. 2A**). There were no other significant differences in PC values (**Fig. 2B**), so subsequent analyses focused on patients with MC1.

**Figure 2.**
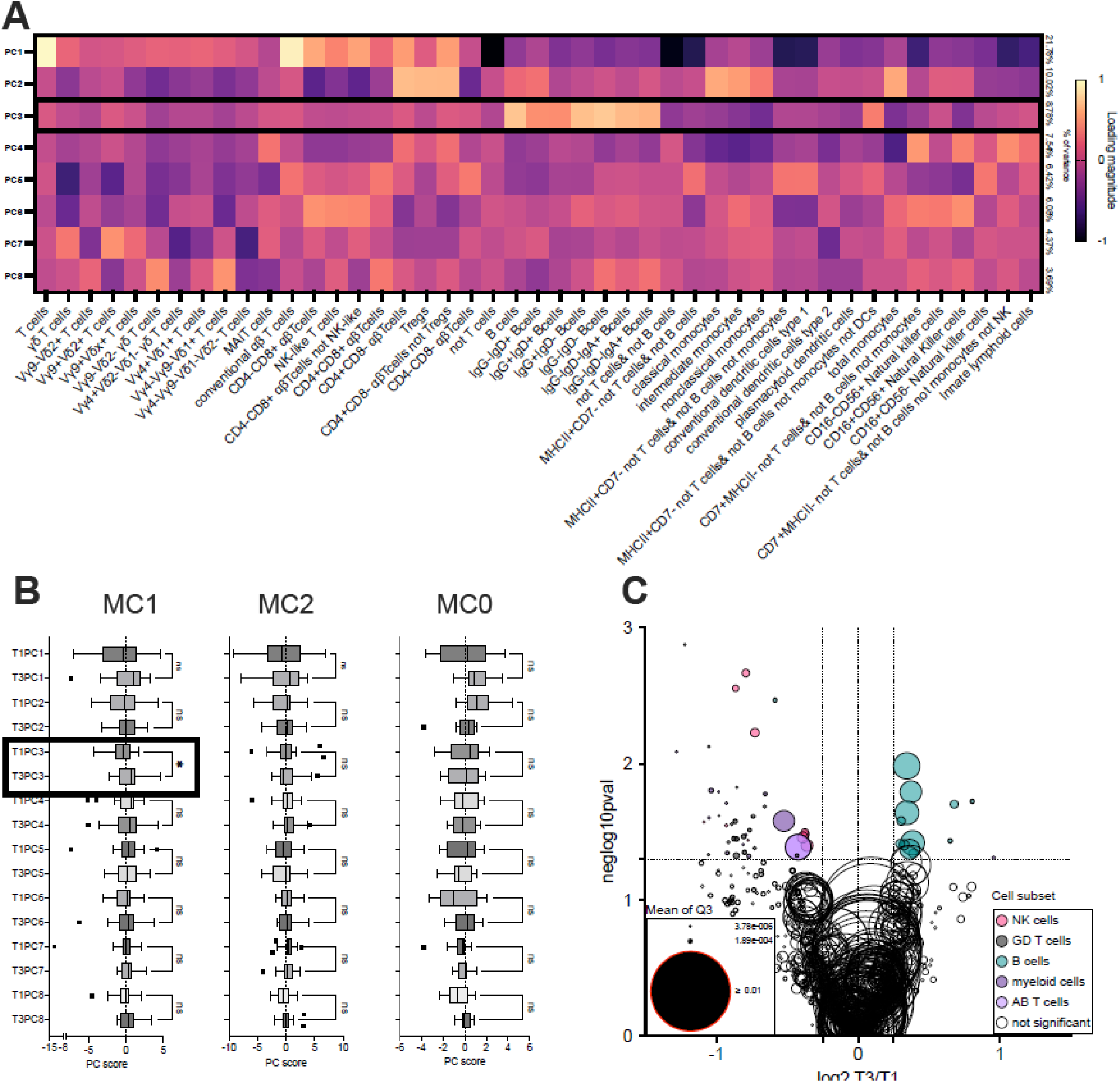
Flow cytometry-based immunophenotyping of CLBP patients’ blood. (**A**) PCA loading matrix of 8 principal components (PCs) by 45 lineage subsets of immune cells determined by multicolour flow cytometry. The loading magnitude is depicted by colour. The percentage of total variance explained b each PC is described on the right side of the loading matrix. (**B**) Comparison of PC scores for individual PCs between MC1 (**B-left**), MC2 (**B-middle**) and MC0 (**B-right**) in PA-high (tertile 3- “T3”) and PA-low (tertile 1 – “T1”) patients. (**C**) Comparison of the proportions of 1021 immune cell populations between MPA-high (T3) and PA-low (T1) patients using a generalised linear mixed effect model. The size of a symbol corresponds to the estimated size of the population in PA-High. The colour of the symbol represents the parent population. Dotted horizontal line represents negative log10 pval=1.3 (pval=0.05). The area between dotted vertical lines represents log2 fold change >-0.25 and <0.25.

The 1021 analysed cell populations were assigned into six major populations: B cells (127 subpopulations), all T cells (31 subpopulations), αβ T cells (276 subpopulations), γδ T cells (219 subpopulations), myeloid cells (213 subpopulations), NK cells (122 subpopulations), and ILCs (31 subpopulations) and 3 populations of mixed phenotypes (**Tab. S3**). Comparisons of all immune cell populations between PA-low and PA-high in MC1 patients revealed an increased proportion of 13 immune cell subsets (**Fig. 2C**, **Tab. 3, Tab. S3**) and a decreased proportion of 43 populations (**Fig. 2C, Tab. S3**). 12 of the 13 populations enriched in PA-high were B cells. Fisher’s exact test confirmed significant enrichment of B cell subpopulations among populations enriched in PA-high patients (p<0.001).

**Table 3.**
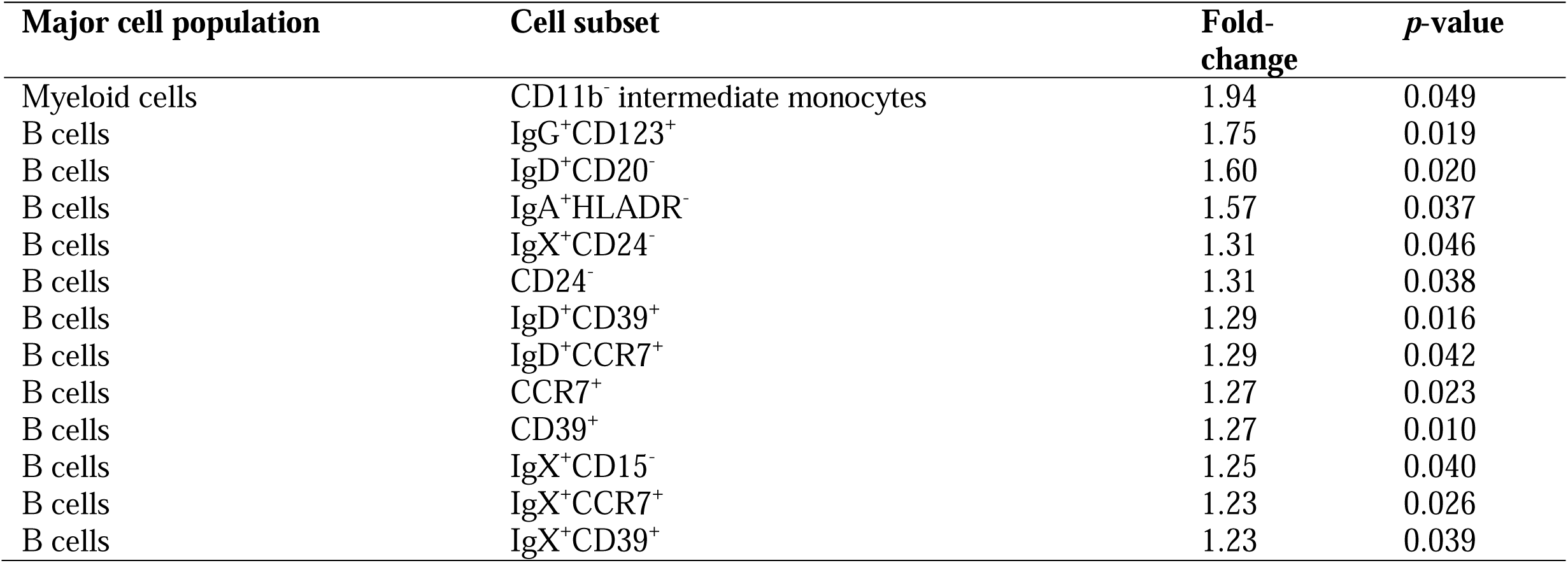
Differentially abundant cell subsets between patients with “PA-high” vs. “PA-low” and MC1.

Enrichment of specific B cell subsets in patients with MC1 and PA-high led us to investigate whether the degree of enrichment correlates with intradiscal PA levels, and if a similar enrichment is observed in patients with MC2 and MC0. There were no significant differences in the proportion of B cells between patients in each PA tertile for all MC types (**Fig. 3A**). Equivalent analysis of CD39^+^ B cells (**Fig. 3B**) and CCR7^+^ B cells (**Fig. 3C**) confirmed the proportions of these cell subsets were significantly higher amongst patients with MC1 and PA-high vs. PA-low, but not different amongst patients with MC2 or MC0. In addition, B cell activation markers were increased in patients with higher intradiscal PA levels. Specifically, there was a significant linear trend for an increase in proportions of CD39^+^ B cells (p=0.009, slope=0.0036) and proportions of CCR7^+^ B cells (p = 0.018, slope = 0.0029) with increasing PA level (from PA-low to PA-high) in patients with MC1. Comparing CD39 expression on CCR7^+^ and CCR7^-^ B cells and CCR7 expression on CD39^+^ and CD39^-^ B cells revealed that B cells expressing either of these markers also expressed significantly higher levels of the other marker (**Fig. 3D**), indicating that they are expressed by the same B cell population. Consequently, the proportion of CCR7^+^CD39^+^ B cells was markedly different between patients with MC1 and PA-high vs. PA-low (**Fig. 3E**). Lastly, we found no difference in median fluorescence intensity (MFI) of CD39 and CCR7 on B cells between PA tertiles, indicating these findings reflect differences in the relative abundance of CCR7^+^CD39^+^ B cells and not differences in the expression levels of those markers (**Fig. 3F**).

**Figure 3.**
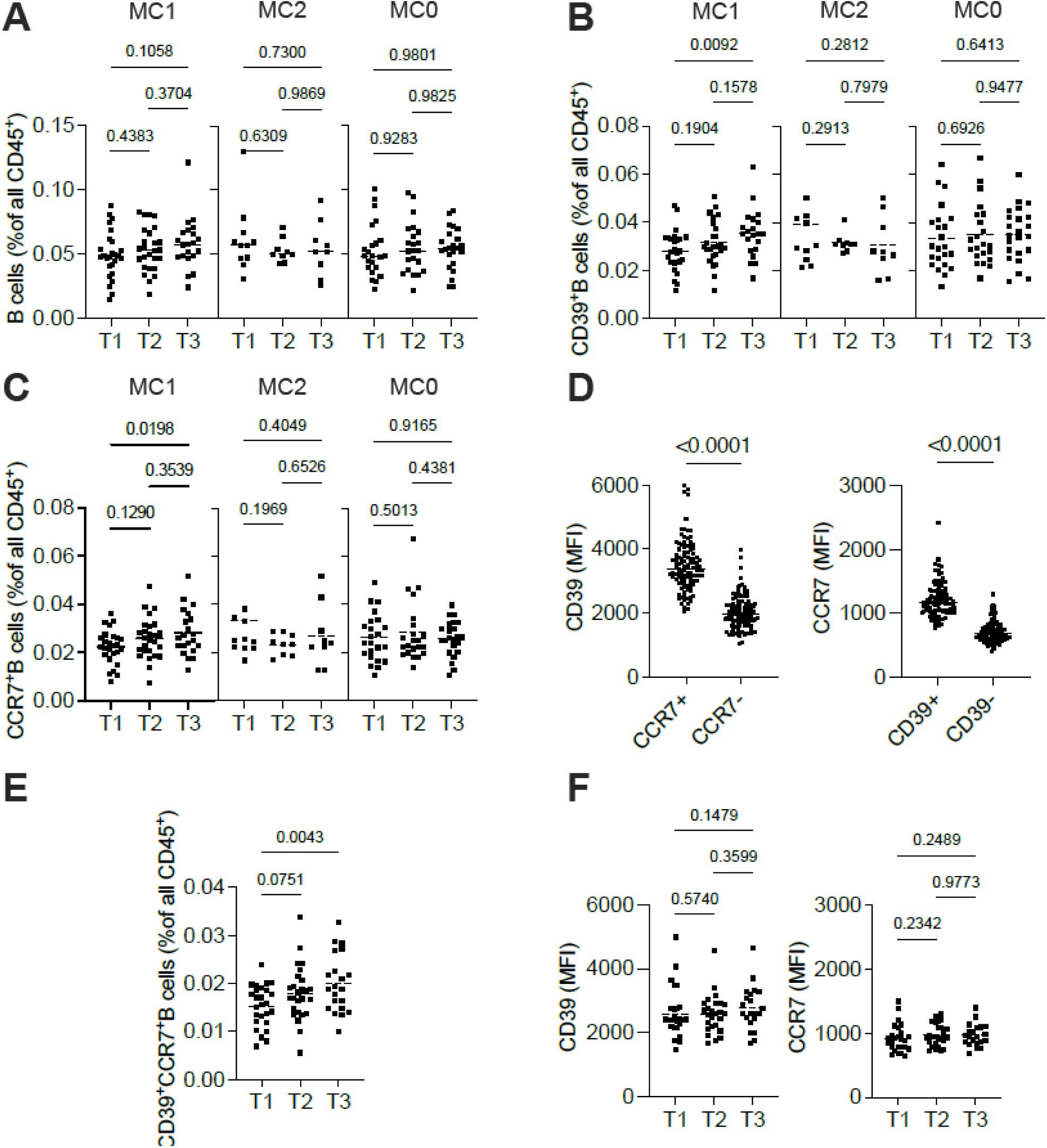
Comparisons of different B cell subsets between patients with PA-low (PT1), intermediate PA (T2) and PA-high (T3) in patients with MC1, MC0, and MC0. **(A)** The proportion of B cells; (**B**) the proportion of CD39^+^B cells; and (**C**) the proportion of CCR7^+^B cells. (**D**) Comparison of CD39 expression on CCR7^+^ and CCR7^-^ B cells (left) and CCR7 expression on CD39^+^ and CD39^-^ B cells (right). (**E**) Comparison of the proportion of CD39^+^CCR7^+^B cells between patients with PA-low (T1), intermediate PA (T2) and PA-high (T3) for patients with MC1. (**F**) Comparison of the median fluorescence intensity of CD39 (left) and CCR7 (right) on B cells from patients with MC1 and PA-low (T1), intermediate PA (T2), and PA-high (T3).

Investigation of the correlation between intradiscal PA values and the proportion of various immune cell subsets revealed significant negative correlations with PA for 44 subsets and significant positive correlations with PA for two subsets (**Fig. 4A, Tab. S4**). CD39^+^ B cells were the most strongly and significantly positively correlated with intradiscal PA levels (ρ =0.274, *p*=0.013), whereas TCM Vγ4δx T cells showed the strongest negative correlation (ρ =-0.351, *p*=0.002) (**Fig. 4A**). Since B cells were the most significantly positively correlated with intradiscal PA levels, we investigated differences within the B cell population using unsupervised clustering by FlowSOM algorithm (**Fig. 4B-D**). Clustering was performed based on the expression of all stained markers relevant for B cells (CCR7, CD11a, CD15, CD19, CD20, CD24, CD25, CD26, CD38, CD39, CD45, CD45RA, CD73, HLA-DR, IgA, IgD, IgG, PD1) (**Fig. 4B-C**). Using a 2-way ANOVA, we observed no overall effect of intradiscal PA level on B cell composition; however, there was a significant enrichment in one specific population, represented by metacluster 3 (**Fig. 4D**). This cluster consisted of IgD^+^ B cells with high surface expression of CD73 (**Fig. 4B**).

**Figure 4.**
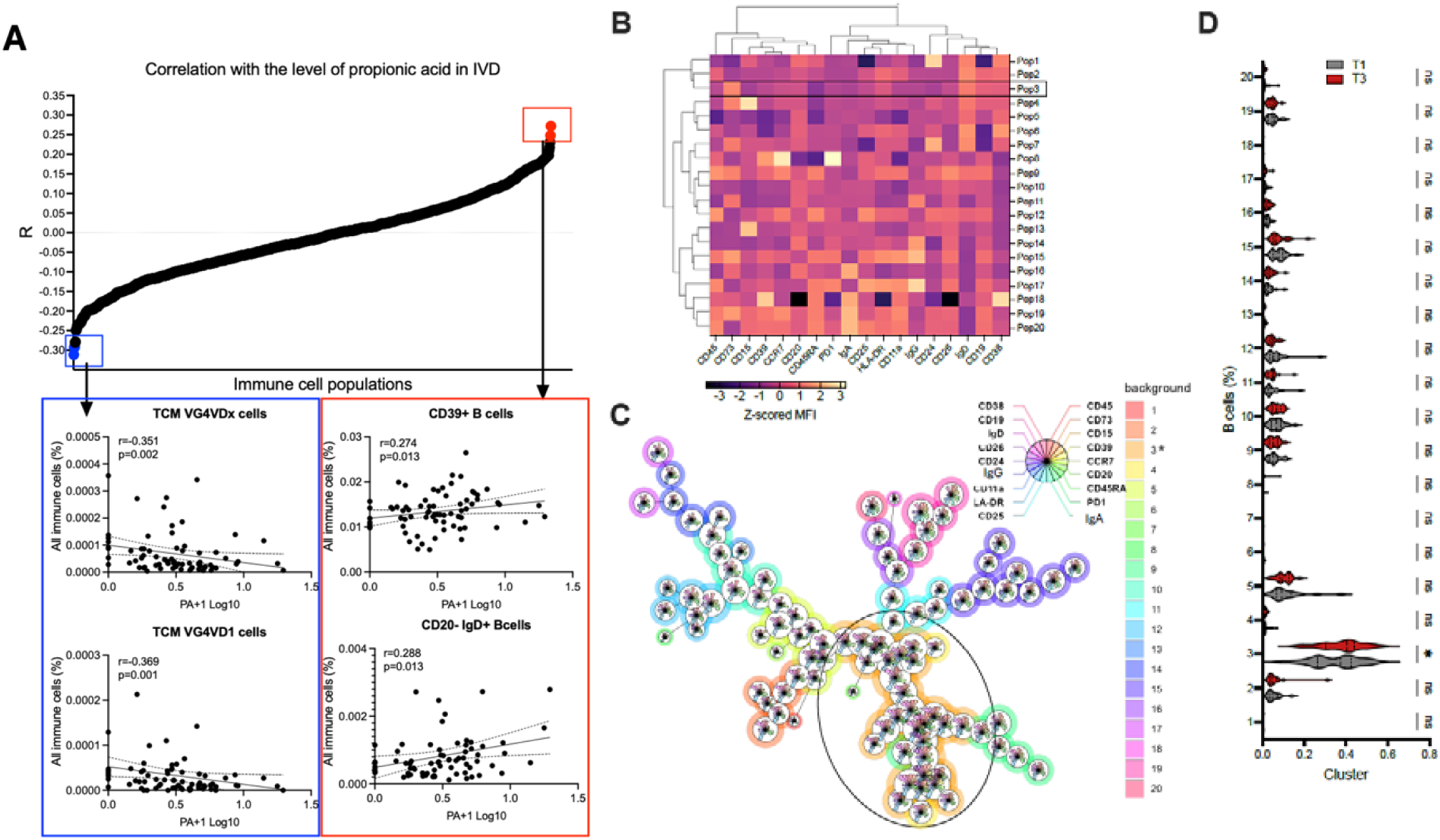
**(A)** Spearman correlation between PA level and immune cell subpopulation in MC1 patients (top). Correlations for the two immune cell populations with the greatest negative Spearman coefficient (blue) and the strongest positive Spearman coefficient (red) are given in the bottom panels. **(B)** Flow Self Organising Map (FlowSOM) clustering of B cells followed by hierarchical clustering into twenty meta-clusters. **(C)** FlowSOM minimal spanning tree visualisation of clustered cytometry data. Each node represents a cluster (SOM node) containing similar cells based on marker expression. The size of the nodes corresponds to the number of cells in the cluster. Edges represent similarity between clusters. Node background colours indicate meta cluster assignment as specified in the legend. Nodes with similar profiles are connected to form a tree structure that preserves local similarity in the high-dimensional space. **D.)** Comparison of FlowSOM meta-clusters using two-way Analysis of Variance revealed similar B cell meta-clusters in patients with PA-high (T3) vs. PA-low (T1). Metacluster 3 was significantly higher in patients with PA-low.

### Serum cytokines

Serum samples from 398 patients were assayed for forty cytokines associated with inflammation. Out of 15,920 total cytokine values, 860 missing values (5.4%) were imputed. Among these were 73 MC1, 29 MC2 and 68 MC0 patients, for whom we also performed MRS-based PA quantification and flow cytometry immunophenotyping. Spearman correlation of blood serum cytokine levels with PA MC1 patients revealed a significant negative correlation between PDGF-BB (ρ =-0.32, p=0.007), TIMP2 (ρ =- 0.31, p=0.008), and RANTES (ρ =-0.23, p=0.049) and PA levels (**Fig. 5A, Tab. S5**). No significant positive correlation was detected. The highest correlation coefficient was measured for IL12-p40 (ρ =0.15, p=0.21). Markedly different results were obtained for MC2 and MC0, with IL-12-p40 and IL-4 showing a significant negative correlation with PA in both (**Fig. S3**). In MC2, a significant negative correlation was also observed with TNFRI, and in MC0 with MCSF and G-CSF (**Fig.S3**). Hierarchical clustering of correlation coefficients of immune cell subsets and PA with cytokine levels in MC1 patients, using Euclidean distance and agglomerative complete-linkage clustering, revealed that correlations of cytokines with PA were closest to correlations of cytokines with B cells and IgD^+^ B cells (**Fig. 5B, Tab. S6**). In MC0 and MC2 patients, PA levels correlate with blood plasma cytokines most closely associated with those of NK cells and nonclassical monocytes (**Fig.S4**).

**Figure 5.**
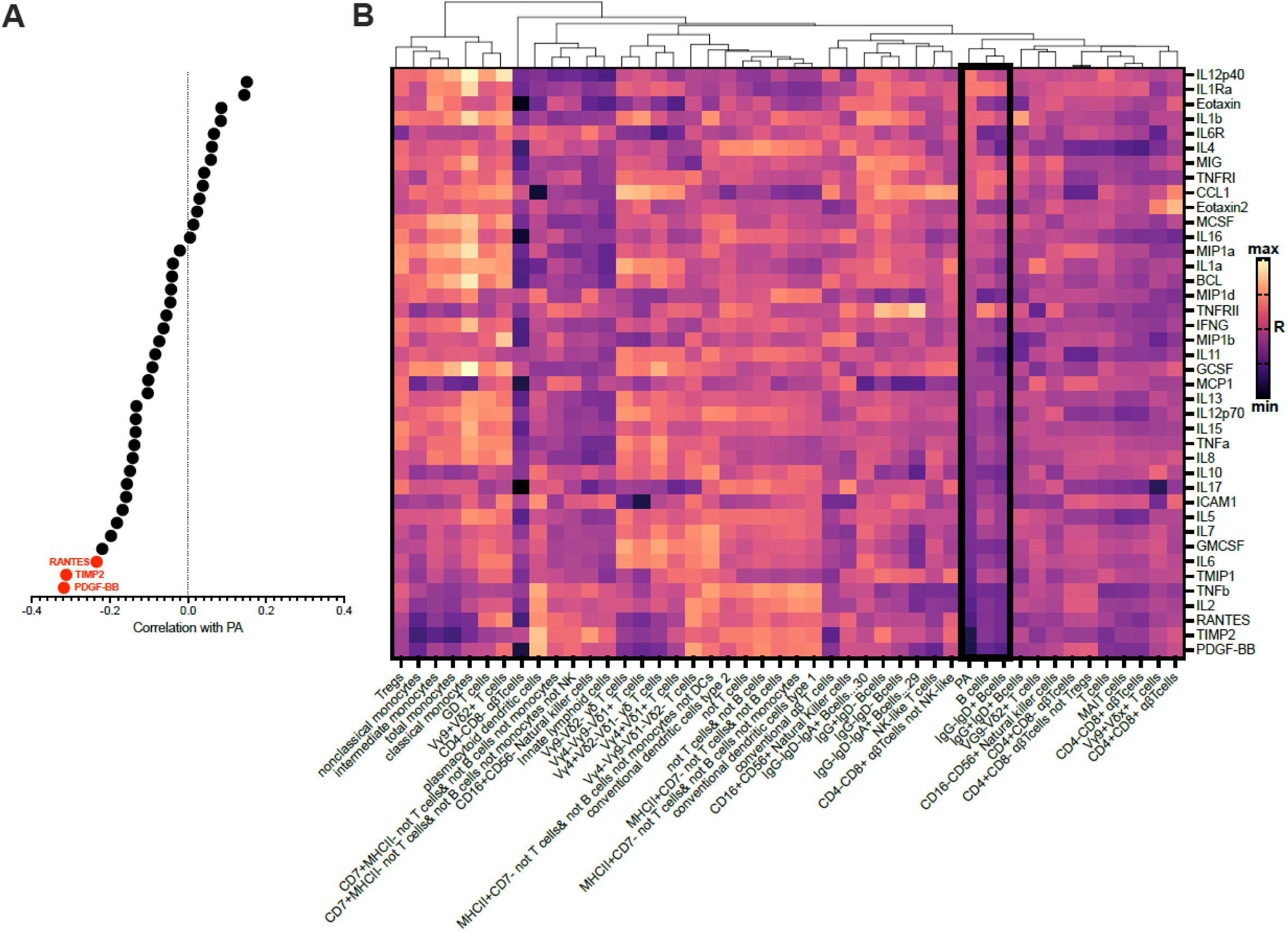
**A.)** Spearman correlation coefficients between PA level and blood serum cytokines. Cytokine with significant correlations are highlighted in red. **B.)** Heatmap depicting Spearman correlation coefficients of major immune cell populations and intradiscal PA with blood plasma cytokines in patients with MC1. Rows are organised based on cytokine correlations with intradiscal PA, from highest to lowest. Columns are organised by hierarchical agglomerative clustering using Euclidean distance and complete linkage. Thick black framing is used to depict the column corresponding to intradiscal PA and the closest clustered immune cell populations – B cells and IgD^+^ B cells.

## 4. DISCUSSION

This study provides convergent evidence from transcriptomic, multiparametric flow cytometry, and serum cytokine analysis showing that intradiscal PA levels are associated with both the relative abundance and activation state of peripheral B cells in patients with MC1, the MC subtype most consistently associated with pain^30^. Notably, these B cell responses were not observed in patients with MC2 or in those without MC, indicating specificity for the MC1 phenotype.

### Biological and Clinical Implications

Our transcriptome analysis revealed pronounced enrichment of gene sets related to adaptive immune responses and B cell activation in PA-high MC1 cases, including upregulation of immunoglobulin heavy chain transcripts and pathways such as “positive regulation of B cell activation” and “B cell receptor signalling”. Flow cytometry further demonstrated that amongst the more than one thousand immune subsets analysed, B cells – particularly those expressing activation markers CCR7 and CD39, and most significantly the CCR7□CD39□double-positive subset – were enriched in PA-high MC1. Data-driven unsupervised clustering independently highlighted an IgD□CD73□B cell metacluster enriched in these patients. These findings indicate that both hypothesis-driven and data-driven strategies consistently converge on B cells as the central immune cell population associated with increased intradiscal PA in MC1.

Cytokine profiling showed that in MC1 patients, PA levels were inversely correlated with serum concentrations of PDGF-BB, TIMP2, and RANTES – factors involved in tissue repair, extracellular matrix modulation, and immune cell recruitment^31–33^. Interestingly, the pattern of PA correlation with cytokines was markedly different in MC1 patients than in MC2 and MC0. In addition, in MC1, hierarchical clustering aligned PA-cytokine correlation patterns closely with cytokine correlation with B and IgD□B cells, which was not observed for MC2 and MC0 patients. These patterns together add a further layer of support to the association of B cells with intradiscal PA levels in MC1 patients.

### Mechanistic Connections Between Propionic Acid and B Cells

The observed association between PA and B cell profiles might reflect: i.) the direct effects of PA on B cells; and/or ii.) the adaptive immune response to *C. acnes* as a PA source.

#### i.) PA as an immunomodulator

PA has been shown to modulate both systemic and tissue-resident immunity by directly influencing B cell function and phenotype^34^. In a murine model and in vitro cultures of human B cells, PA can stabilise IL-10 expression in B cells, promoting a regulatory phenotype and supporting immune tolerance^34^. PA can also suppress activation of the toll-like-receptor (TLR)4/NF-KB pathway and has a rather regenerative than pro-inflammatory effect^35,36^. This might be relevant in MC1 pathobiology because discs at MC1 levels contain more TLR4 agonists^10^. However, the effects of PA are highly context- and cell-type-dependent, and PA can act as both a regulator and a potentiator of inflammation^37^. PA, when released in high concentrations during bacterial fermentation in lipid-rich, hypoxic environments, strongly inhibits HDAC8 and HDAC9 in human epithelial keratinocytes, leading to increased histone acetylation at the promoters of inflammatory genes. This epigenetic mechanism substantially enhances the expression of proinflammatory cytokines (such as IL-6, IL-8, and CCL5/RANTES) in keratinocytes following TLR stimulation, contrasting the anti-inflammatory effects of HDAC inhibition in monocytes and other immune cells^37^. Thus, while the precise role of PA in MC1 remains to be revealed, our present findings motivate future investigations about the effects of PA on intervertebral disc cells and cells of the cartilage endplate.

#### ii.) Immune response to *Cutibacterium acnes* as a PA source

An alternative explanation is that PA serves primarily as a surrogate marker for *C. acnes* presence and activity in the disc. *C. acnes* has been consistently found in intervertebral discs, with higher prevalence or concentration in MC1, and it is known to produce PA as a major metabolic byproduct^38,39^. In this model, elevated PA, as measured by MRS, reflects a higher bacterial burden, and the observed enrichment of B cell subsets in PA-high MC1 patients represents an adaptive immune response against bacterial antigens. Whether such B cell responses are protective by contributing to bacterial clearance, or pathogenic by sustaining inflammation and pain, remains unresolved. The model of immune response against *C. acnes* is further supported by the transcriptomic signature of “Defense response to bacterium” and of activated neutrophils in PA-high in MC1. *C. acnes* has been shown to trigger inflammation in discs, and neutrophils were activated in MC1 bone marrow adjacent to discs with high *C. acnes* loads^11,40^.

We observe that the B cells accumulating in PA-high MC1 patients are often CCR7□, CD39□and CD73□, which suggests their activation^41,42^. However, the expression of CD39 and CD73 can also be associated with the immunomodulatory effects of PA. CD39 and CD73 are key ectonucleotidases involved in the generation of extracellular adenosine, a molecule with anti-inflammatory properties^43,44^. Recent studies, for example, show that PA exposure enhances the suppressive capacity of regulatory B cells and increases IL-10 and TGF-β expression, linking microbial metabolite sensing to the control of autoimmunity and inflammation^34^. These immunoregulatory B cell subsets have been implicated in a variety of chronic inflammatory and autoimmune diseases^45^. It thus remains to be determined whether the B cell accumulating in MC1 are primarily protective or pathogenic.

### Limitations of the Study

The main limitation of this observational study is that disc tissues were unavailable for analysis. Thus, we did not measure the amount of intradiscal *C.acnes* or directly validate the concentration of intradiscal PA. We previously found that *C.acnes-* and PA-injected bovine discs had clear MRS peaks corresponding to the chemical shift of PA (∼1.05 ppm), whereas un-injected discs lacked the characteristic peak^17^. Also, the signal intensity at this characteristic chemical shift increased with increasing volume of *C.acnes* in the injectate. While these findings support the sensitivity and specificity of MRS, we cannot exclude the possibility that other sources of PA contributed to the MRS signal in this study. Furthermore, we performed an associative analysis, which limits interpretation due to the possibility of confounding and selection bias.

An additional limitation of the study is that differences in circulating inflammatory factors or cell populations may reflect underlying comorbidities unrelated to cLBP status. Patients were excluded from the study if they were pregnant or had known conditions that affect systemic immune markers, e.g., ankylosing spondylitis, rheumatoid arthritis, polymyalgia rheumatica, psoriatic arthritis, lupus, cancer, discitis, or osteomyelitis^46^. However, we cannot exclude the possibility that some patients had undiagnosed conditions or infections that influenced the composition and activation of the immune system.

### Translational Relevance and Future Directions

Identifying B cells as central immunological mediators in MC1 raises the possibility of repurposing or developing B cell-directed therapies for patients with MC1-related back pain and PA-high, mirroring advances made in other immune-mediated musculoskeletal diseases such as rheumatoid arthritis^47–49^. The observed B cell signature may also serve as a biomarker for *C. acnes*-caused MC1 and to stratify MC1 patients for antibiotic therapy, in case future studies can link high disc PA to high *C. acnes* loads. Further mechanistic studies are warranted to elucidate the antigenic specificity and functional potential of these B cell subsets, as well as to develop experimental models that directly test the modulatory role of PA in chronic discogenic pain.

## Conclusion

This study establishes B cell activation as a core immune correlate of high intradiscal PA in LBP patients with lumbar MC1. Our findings indicate that local PA changes in the disc are linked to systemically detectable changes of the adaptive immune system in MC1 patients, suggesting that MC1 has a systemic immune component. This supports our recent findings of HLA association with MC1. These findings advance our understanding of the immunometabolic interface in spine disease and support a personalised biomarker- and cell-targeted therapeutic approach for MC1-related LBP.

## Data Availability

All data produced in the present study are available upon reasonable request to the authors

## ACKNOWLEDGEMENTS

This work was funded by National Institutes of Health grants U19 AR076737, UH2 AR076719, UH3 AR076719 and P30 AR075055, the Swiss National Science Foundation (207989), and the Promedica Foundation (1667/M). We acknowledge the Swiss Center for Musculoskeletal Biobanking at the Balgrist Campus, the Functional Genomic Center Zurich, and Kenta Robin Brender for their support in transcriptome analysis. This content is solely the responsibility of the authors and does not necessarily represent the official views of the National Institutes of Health.

The Back Pain Consortium (BACPAC) Research Program is administered by the National Institute of Arthritis and Musculoskeletal and Skin Diseases (NIAMS).

The REACH Investigators would like to express their gratitude to the members of the comeBACK clinical site research team, especially the Clinical Research Coordinators (CRCs) for their commitment and contributions toward the successful conduct of the study (in alphabetical order): Jamie Ahn^3^, Kristina Benirschke^1^, Alexandra Bryson^1^, Katherine Bunda^4^, Briana Davis^1^, Carolina Dorofeyev^2^, Rosalee Espiritu^4^, Pirooz Fereydouni^1^, Aamna Haq^1^, Nicholas Harris^1^, Sara Honardoost^3^, Gabriel Johnson^1^, Jennifer Johnson^1^, Edward Lingayo, Jr^2^, Robert Miller^3^, Phirum Nguyen^4^, Christopher Orozco^1^, Lindsay Ruiz-Graham^2^, Kie Shidara^1^, Kaitlyn Smith^1^, John (Boyuan) Xiao^1^, Michelle Yang^1^

CRC Affiliations

1University of California, San Francisco

2University of California, Davis

3University of California, Irvine

4University of California, San Diego

**\*REACH Investigators**

The Core Center of Patient-centric, Mechanistic Phenotyping in Chronic Low Back (REACH) investigators include the following University of California, San Francisco (unless noted otherwise) personnel in alphabetical order:

Zehra Akkaya, MD

Prakruthi Amarkumar, PhD

Jeannie Bailey, PhD

Julia Barylak Sigurd Berven, MD

Andrew Bishara, MD

Dennis M. Black, PhD

Noah Bonnheim, PhD

Atul Butte, MD, PhD

Joel Castellanos, MD (University of California, San Diego)

Jennifer Cummings

Karina Del Rosario, MD

Emilia Demarchis, MD

Sibel Demir-Deviren, MD

Susan K. Ewing, MS

Adam R. Ferguson, PhD

Aaron Fields, PhD

Scott M. Fishman, MD (University of California, Davis)

Sergio Garcia Guerra

Fatemeh Gholi Zadeh Kharrat, PhD

Xiaojie (Summer) Guo

Misung Han, PhD Trisha Hue, PhD

J. Russell Huie, PhD

C. Anthony Hunt, PhD

Anastasia Keller, PhD

Karim Khattab

Roland Krug, PhD

Gregorji Kurillo, PhD

Feng Lin

Thomas M. Link, MD, PhD

Jeffrey Lotz, PhD

John Lynch, PhD

Tong Lyu

Rob Matthew, PhD

Wolf Mehling, MD

Esmeralda Mendoza, MPH

Praveen Mummaneni, MD, MBA

Caroline Navy

Conor O’Neill, MD

Jessica Ornowski

Thomas Peterson, PhD

Ananya Rupanagunta (University of California, Berkeley)

Aaron Scheffler, PhD, MS

Shalini Shah, MD (University of California, Irvine)

Irina Strigo, PhD

Naoki Takegami, MD

Abel Torres-Espin, PhD (University of Waterloo)

Salvatore Torrisi, PhD

Sachin Umrao, PhD

Rohit Vashisht, PhD

Joanna Veres

An (Joseph) Vu, PhD

Mark Steven Wallace, MD (University of California, San Diego)

Lucy Ann Wu, MPH

Po-Hung Wu, PhD

Fadel Zeidan, PhD (University of California, San Diego)

Patricia Zheng, MD

Jiamin Zhou, MS

---

**Figure S1.**
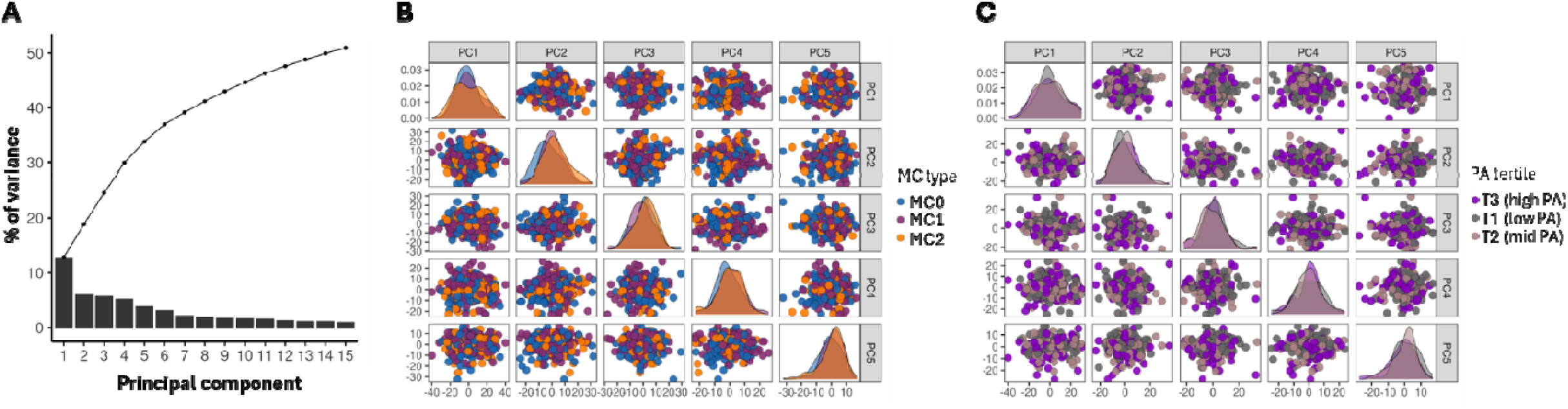
Whole blood bulk-RNA sequencing analysis from CLBP patients. (**A**) Scree plot of principal component analysis (PCA) indicating percentage of total variance explained by each PC (bars) and cumulative (line). (**B**) Paired plots of the first five principal components (PC) colored by Modic change (MC) type and (**C**) by PA tertile.

**Figure S2.**
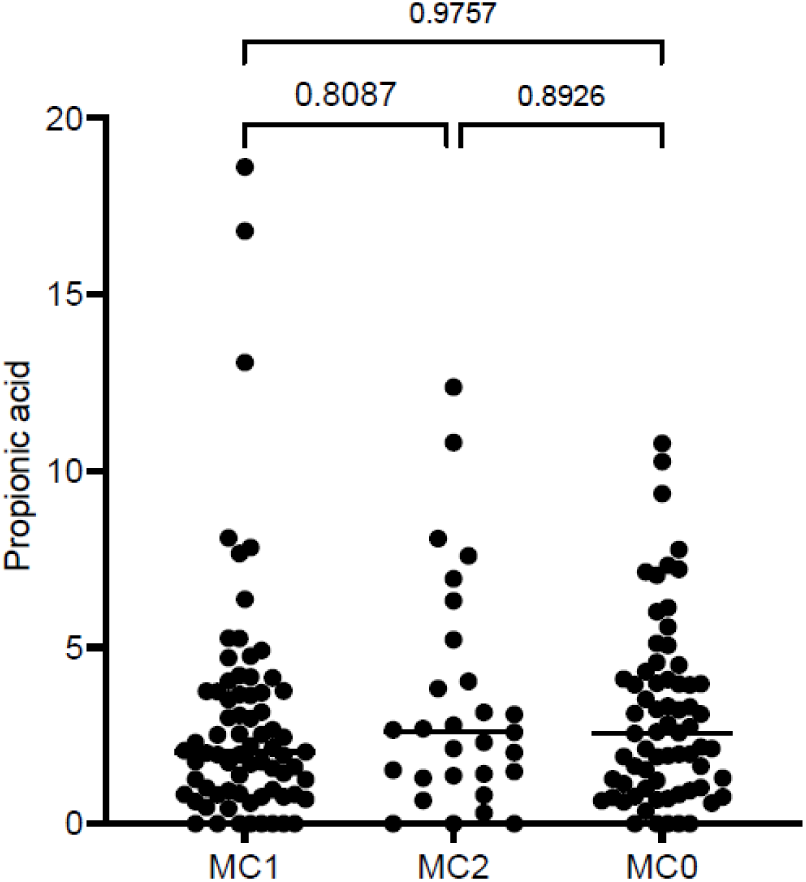
Comparison of intradiscal PA levels among MC1, MC2 and MC0 CLBP patients.

**Figure S3.**
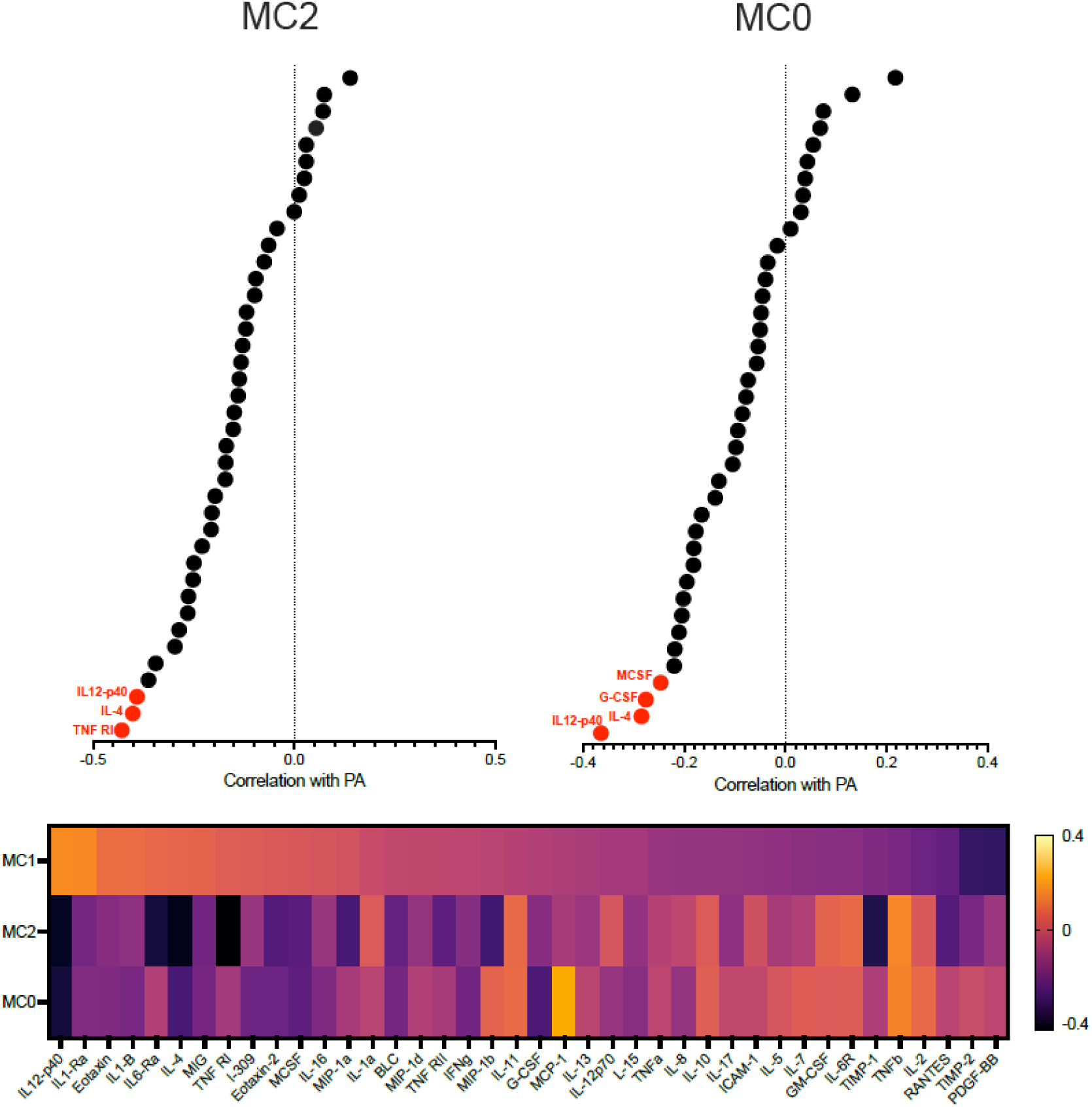
**A.)** Spearman correlation coefficients between PA level and blood serum cytokines in MC2 and MC0 patients. Cytokines with significant correlations are highlighted in red.

**Figure S4.**
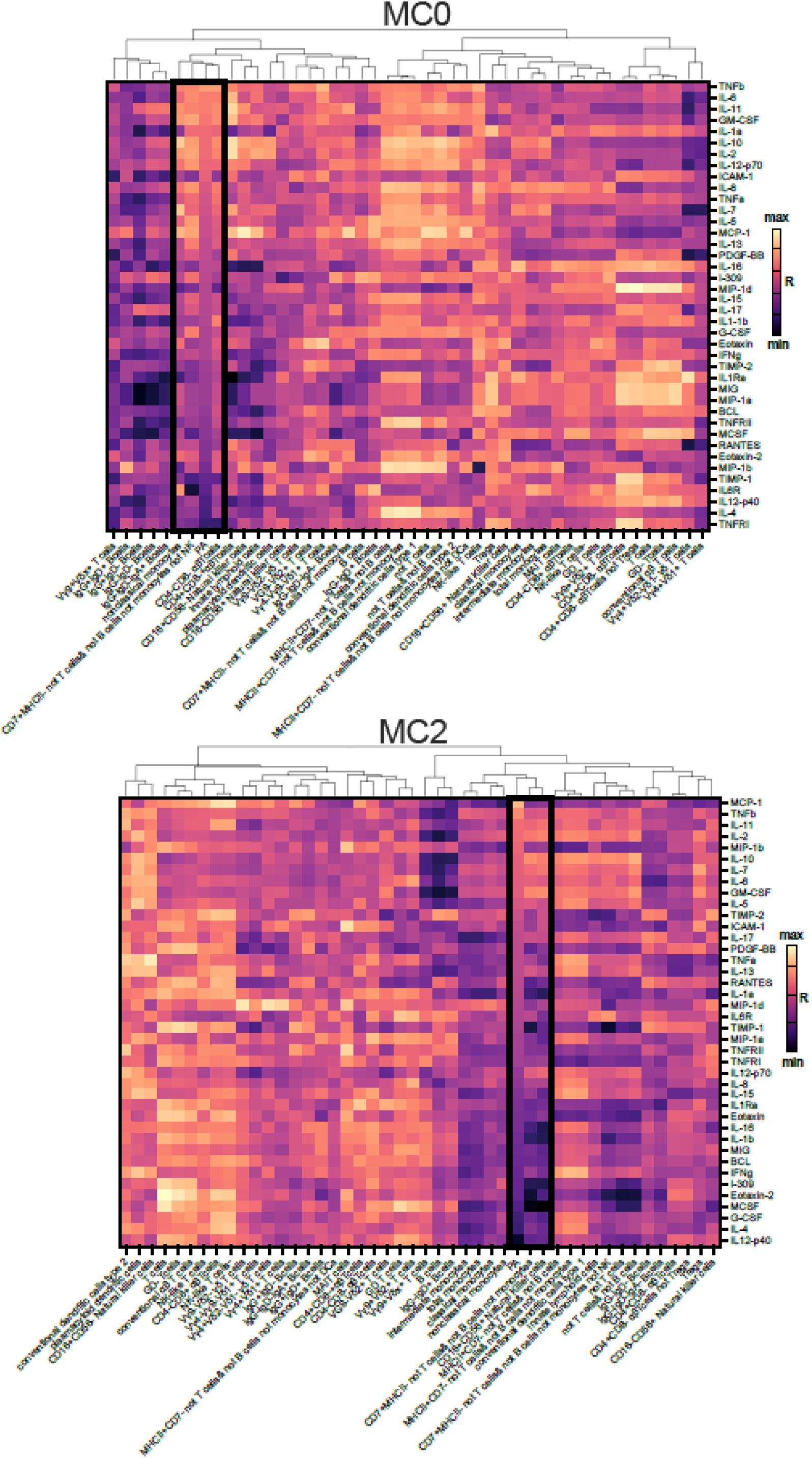
Heatmap depicting Spearman correlation coefficients between blood serum cytokines and major immune cell populations and intradiscal PA in MC2 and MC0 patients. Rows are organised based on cytokine correlations with intradiscal PA, from highest to lowest. Columns are organised b hierarchical clustering. Black framing is used to depict the column corresponding to intradiscal PA and the closest clustered immune cell populations – B cells and IgD^+^ B cells.

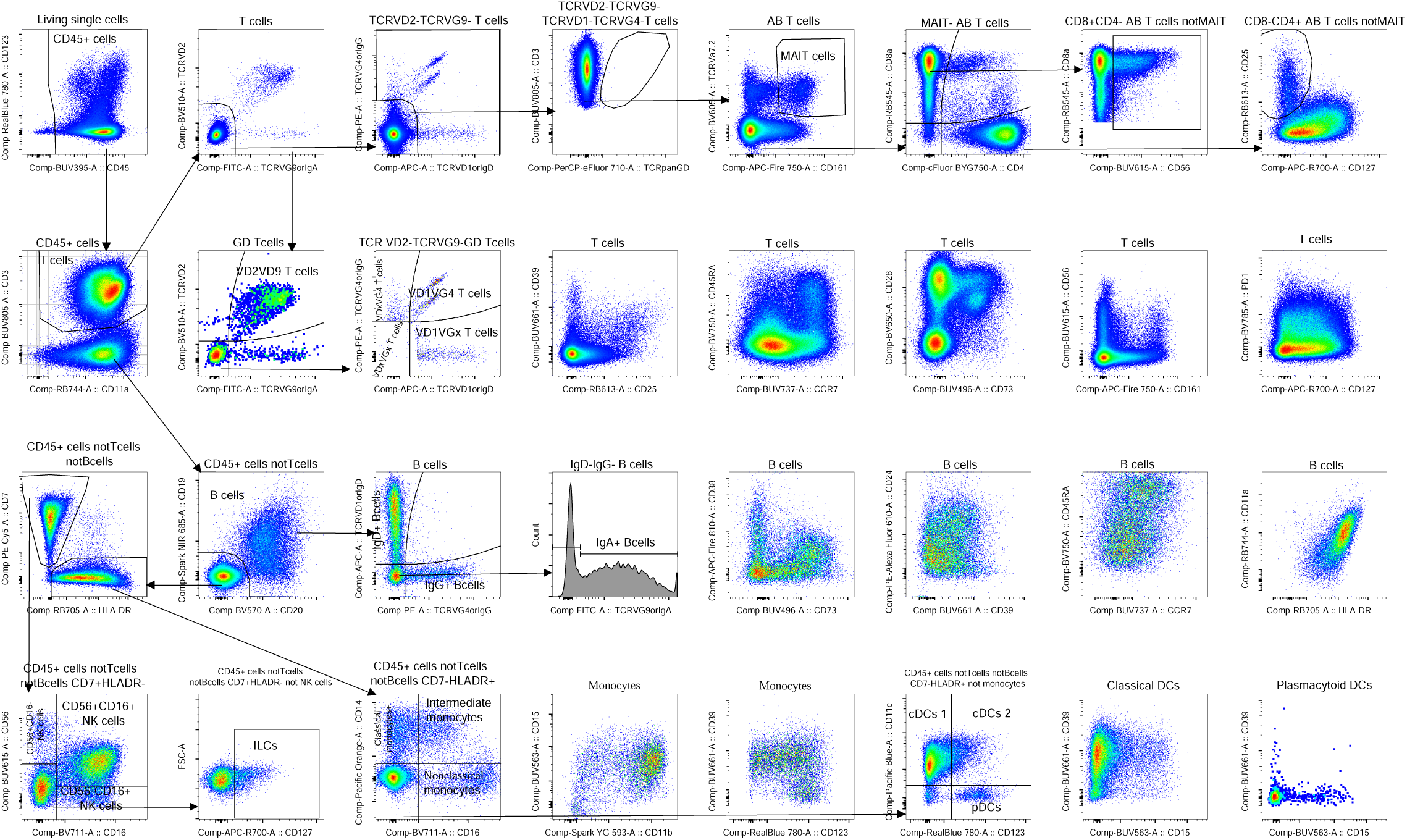

